# Green Heart Louisville: Intra-urban, hyperlocal land-use regression modeling of ultrafine particles

**DOI:** 10.1101/2023.03.03.23286768

**Authors:** Pradeep Prathibha, Raymond Yeager, Aruni Bhatnagar, Jay Turner

## Abstract

Exposure to ultrafine particles (UFP) is increasingly linked to adverse health outcomes. While nation-wide air monitoring networks in the United States do not measure UFP, small-scale measurements have revealed persistent patterns in urban UFP. This project maps hyperlocal UFP in a 12 km^2^ study area of a health effects study in Louisville, KY, through mobile measurements to elucidate the relationship between the urban landscape and UFP exposures. We measured UFP number concentration along all drivable streets (∼340 km) during daytime and nighttime on both weekdays and weekends. After deconvoluting UFP levels to isolate local signals from neighborhood and urban signals, we fitted a land-use regression (LUR) model to explain differences in local UFP as a function of characteristics of the built and natural environment.

Median UFP in the study domain was 6,850 #/cm^3^, which is comparable to urban background measured or estimated for other U.S. cities. UFP was higher during the weekend than on weekdays, potentially due to changes in local activity (e.g. increased restaurant hours) apparent at fine spatial scales. The final LUR model explained 61% of the spatial heterogeneity in log(UFP). Leave-one-area-out cross validation revealed overprediction in regions farther from highways and underprediction in regions with dense food service locations and major roads. This suggests that additional mobile measurements to capture longer-term, robust UFP may yield improved models.

## Introduction

*Note: “UFP concentration” refers to particle number concentrations unless specified otherwise*.

Recent epidemiological and toxicological studies have demonstrated increased health risks from exposure to ultrafine particles (UFP, particulate matter with aerodynamic diameter <100 nm) than those in higher size ranges (1). UFP can penetrate the most distal conductive pathways in the lungs, leading to systemic circulation and toxicity (2,3). Toxicological studies to date suggest that in addition to the cardiopulmonary effects associated with exposure to fine particulate matter, exposure to UFP is associated with increased markers of brain inflammation (4). Despite the absence of long-term epidemiological assessments of UFP exposure, evidence of enhanced toxicity compels characterizations of exposure.

In most environments, motor vehicle emissions are the principal source of UFP (5). Urban UFP levels are estimated to have diminished by nearly 30% in the past two decades due to regulatory measures implemented to reduce fine particulate mass concentrations (6). Nevertheless, UFP number concentrations in near-road urban environments can be up to 18-fold elevated relative to those in rural areas (7). Finer-scale assessments in recent years have also determined that solid fuel combustion (coal-fired power plants, biomass burning) and cooking also emit UFP, albeit with more heterogeneous composition than those from automotive emissions (8).

Despite the amassing toxicological evidence of adverse health outcomes from exposures to UFP, this pollutant is not monitored in national ambient air quality monitoring networks in the United States. In response, investigators have conducted small-area UFP characterizations through fixed and mobile measurements. In one of the earlier characterizations of UFP exposures, Aalto *et al*. showed that UFP levels in European cities exhibited spatiotemporal differences: winter concentrations were twice as high and summer concentrations were half as much as the mean annual concentrations in Stockholm, whereas in Rome and Barcelona, winter concentrations were 4-10-fold higher than those from the summer (9). This study employed a fixed-site study design where six condensation particle counters contemporaneously measured UFP across six cities. Assessments since then have been far more sophisticated, often combining fixed-site network and mobile measurements, and have detected persistent urban- and residential-scale patterns in UFP exposures (10). The subsequent modeling using land-use regression (LUR) has demonstrated modest to good (0.3 < adjusted R^2^ < 0.6) capacity to explain total variation in UFP in intra-urban to nation-wide scales (11–16). However, systematic reviews have found that the accompanying epidemiological evidence for health effects is either insufficient or inconclusive (3–5).

This work aims to characterize UFP hyperlocal heterogeneity to inform small-area assessments of health effects. We conducted mobile measurements across seasons between fall 2018 through early spring 2020 to characterize UFP in Louisville, KY, and constructed LUR models to quantify the contribution of the land use characteristics to UFP spatial heterogeneity.

## Methods

### Study area

The mobile measurements were conducted in a 12 km^2^ study domain located ∼5 km east of the Ohio River and with a population of 25,000 in metropolitan Louisville (38° 11’ 38”, -85° 46’ 29”). The study area (Figure 1), chosen for a prospective cohort study of cardiovascular disease risk and neighborhood level greenness (Green Heart Louisville), is bisected from east to west by a major interstate highway (I-264, Watterson Expressway), flanked along the northwest and east by freight railway lines, and is adjacent to an international airport that serves as the worldwide cargo hub of a major shipping company. While land use within the study domain is mostly residential with some commercial spaces along major roads, land use outside the perimeter is mostly commercial or industrial.

**Figure 1.**
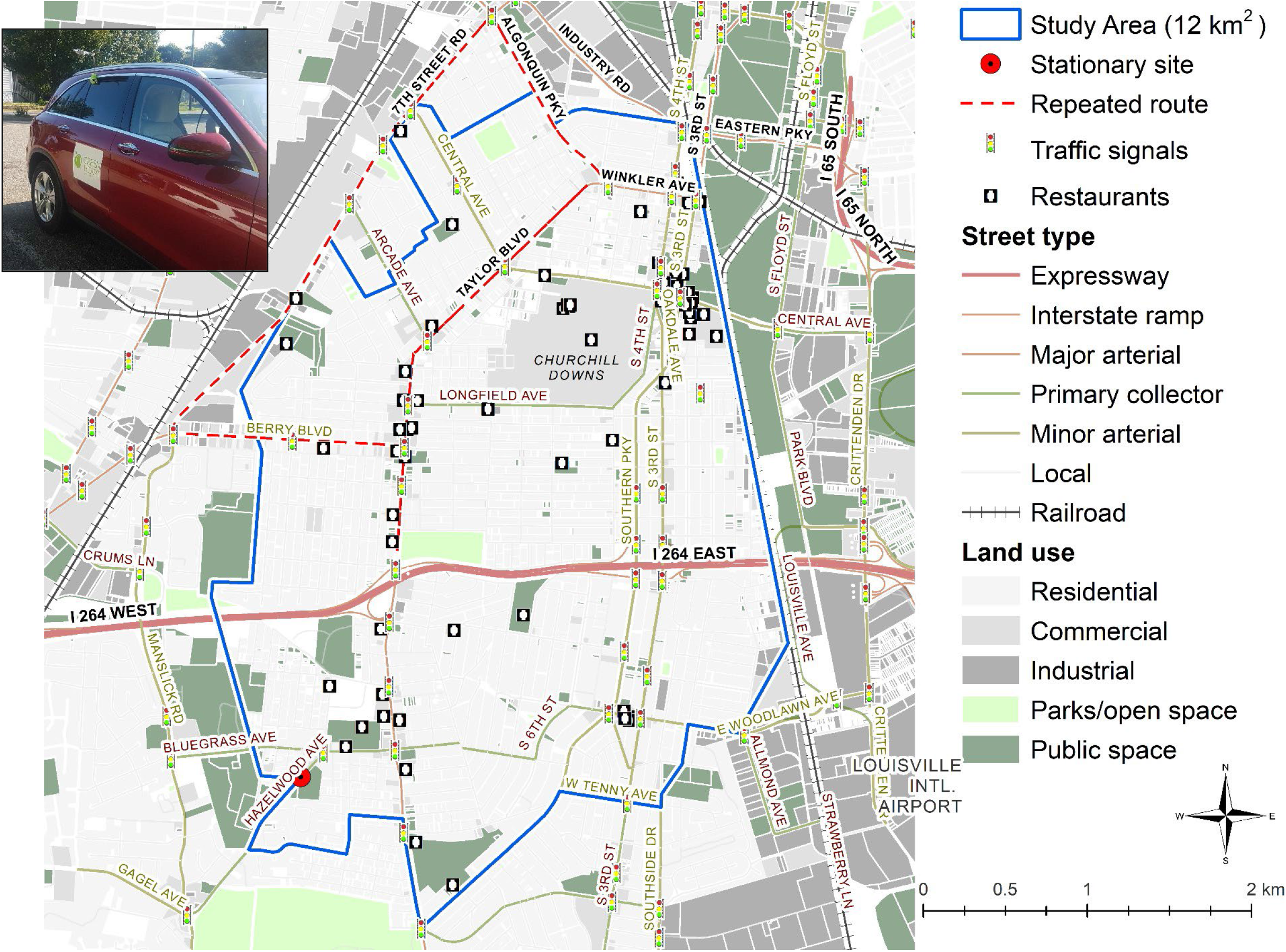
Land use within the study domain (12 km^2^) where UFP number concentrations were mapped using a gasoline-powered vehicle (inset).

### Measurement

Most assessments—including the present work—assume that in typical environments, UFP dominate fine particle number concentrations (PNC) and thus, can be robustly characterized by particle counters rather than more sophisticated and more expensive instrumentation to measure UFP mass concentrations (6,7). We used three fast-response (1 Hz) mixing condensation particle counters (MCPC, Brechtel Inc., Hayward, CA, range: 8-2000 nm particles, η = 50% at 8 nm) to measure UFP number concentrations: two were collocated (for redundancy) in a gasoline-powered car with a BT-Q1000XT global positioning system (GPS, Qstarz International Co. Ltd., Taiwan, R.O.C.) logging the location every second, while the third was set up at a stationary site. All instruments were downstream of 2.5-μm size cut cyclone. In the vehicle, the inlet was positioned ∼10 cm above the roof of the car to avoid the flow reattachment zone and faced perpendicularly away from the cabin to minimize over- or under-sampling. The cyclone was connected to the MCPCs inside the vehicle using ∼1.2 m of steel tubing, and this set up (length, bends, and material) was replicated for the conduit at the stationary site so that wall losses would be comparable. Note that the stationary site is not intended to be in a background location, but rather, to provide fixed-site data for deconvoluting larger-scale signals (see next section).

#### A note on the mobile platform

Electric vehicles were deemed unnecessary for this work as recent studies have shown limited to no interference in measured data due to exhaust from gasoline-powered mobile laboratories. Tailwind speeds need to be greater than the vehicle speed for self-sampling to be a concern, and Tan *et al*. determined that pure tail winds occur for at most 7% of the monitoring duration while driving in typical urban landscapes and Robinson *et al*. observed no increase in aerosol mass concentrations when the mobile laboratory was stationary (17,18). While self-sampling was not extensively evaluated in this work, we confirmed that 1-s UFP in a residential street measured while the vehicle was stationary for 20 mins with the engine off and then on (Figure 8, Supplementary material) were not statistically different (p>0.05).

We conducted multi-day campaigns between August 2018 and February 2020, collecting day- and night-time measurements on weekdays and weekends. Each driving session had a 10-min warm up followed by 4-8 hours of mobile measurements and included a repeated route (Figure 1). For every 3 hours of mobile measurements, we collected 15 minutes of collocated data at the stationary site (vehicle parked ∼10 m away with engine off). We classified data collected between 11 PM and 6 AM as nighttime measurements, while daytime measurements could be taken anytime between 8 AM and 8 PM; note that this delineation does not account for the rush hour traffic and changing mixing layer depth. We covered all drivable streets in the study domain at least four times (at least one of which was driven in the opposite direction) at 8-16 kph for each time of day and day of the week combination. Note that this driving speed does not permit complete coverage of the study domain in each day of driving, so to accommodate comparisons of daytime and nighttime UFP, we covered the same neighborhoods of the study domain during the daytime and nighttime measurements within 24 hours. We classified measurements conducted between 11 PM on Fridays to 6 AM on Saturdays as weekday, nighttime data; consistent with this, measurements between 11 PM on Sundays to 6 AM on Mondays were designated as weekend data.

### Data aggregation

First, we combined UFP and GPS data with a one-second delay to account for the lag time due to the inlet tubing residence time (∼0.8 s) and the instrument response time (0.2 s, calculated by the manufacturer). We determined the collocated precision (CP) between the stationary and mobile MCPCs for each driving session and if this value was lower than 95%, a scalar correction was applied to the mobile-platform data. Then, we snapped each GPS location to the nearest 30-m road segment, discretizing the drivable streets in the study domain into ∼8500 features. Each segment was assigned the median of all UFP measurements that were attached to it. More so than other measures of central tendency, the median can provide stable exposure estimates in near-road environments where instantaneous pollutant levels vary rapidly (19). We selected this snapping distance as it was within the GPS error in covered areas (tunnels, heavily canopied streets) and the spatial scale at which previous mobile assessments have detected persistent patterns in urban UFP (20). While data were distinguished by day- and night-time as well as weekday and weekends for group-wise comparisons, measurements were combined for subsequent analyses to achieve the 10-20 drive days per street recommended by previous mobile studies to robustly capture key spatial patterns (19,21). This portion of the work was completed in ArcMap 10.6.1 (Esri Inc., Redlands, CA).

### Predictor variables

Street centerline (2020), railway (2019), land use (2019), food service locations (2021), signalized intersections (2021), and airports (2019) datasets were obtained from the City of Louisville’s open data portal (22). For each of the ∼8500 segments of drivable streets, we calculated proximity (distance from the centroid) to expressways, interstate ramps, primary collectors, major arterials, minor arterial roads, restaurants, airports, and traffic signals. To quantify land use, we created buffers up to 500 m in 50 m increments centered at the centroid of each segment and within each, calculated the area of residential, commercial, industrial, park and open spaces, public spaces, and road by type (classifications mentioned above and local road). Greenness was quantified in each buffer through 1) 7-inch NDVI raster after excluding cells with NDVI < 0.17 (impervious surfaces, NDVI < 0, or grasses 0 < NDVI < 0.17) and 2) as well as tree-specific leaf biomass and leaf area predicted by collaborators. All calculations were completed in ArcMap and generated 148 explanatory variables.

### Deconvolution

As the present analysis focuses on spatial heterogeneity in UFP, we deconvoluted the stationary monitoring data to isolate neighborhood- and urban-scale pollutant levels, then subtracted these from mobile UFP to isolate local signals. This approach was proposed by Brantley *et al*. to distinguish variations in UFP at various time scales: in brief, UFP data is smoothed using a 30 s rolling median, binned in 1 min windows, and fitted with a smooth thin plate regression spline through the local minimum of each spline (23). We repeated this for varying bin widths (2, 3, 5, 6, 10, 15, 20, 30, 45, and 60 min). In each case, the splined minima is subtracted from UFP measured along the repeated route (to leverage the large number of drives (N>40) on these segments; including other streets would have been computationally intensive and prone to bias due to the lower repetitions). The subtraction generated 11 possibilities of local signals. Then, we calculated the Pearson’s correlation coefficient between these median UFP signals and the proximity to roadway parameters (distance to expressways, interstate ramps, primary collectors, major arterials, minor arterial roads). The buffer distances at which the coefficients drastically decreased indicated the zones of representations of these roads. These distances were 150 m and 450 m; assuming an average driving speed of 10 kph, this meant the neighborhood and urban signals shifted every 54 s and 182 s, respectively. Therefore, neighborhood level signals were isolated using splined minima of 1 min windows and urban level signals were isolated using those from 3 min windows.

This method assumes that neighborhood and urban signals are uniform (temporally coupled) within the study domain, and the remaining differences in the measurements are attributable to local variations in emissions and the built environment. Operationally, this assumption allows the use of any location within a study domain as the stationary site to determine neighborhood and urban signals (11). This portion of the analysis was completed in R (version 4.1.1) using the *zoo* and *mgcv* packages.

### Model development and validation

The dependent variable was log-transformed due to strongly asymmetric distribution. We calculated Spearman’s correlation coefficients between UFP and all predictors and built the LUR model through supervised forward stepwise regression, adding independent variables with the highest correlation to UFP one at a time. Predictors were retained if they increased the adjusted R^2^ of the model by ≥1% and discarded if they did not change the R^2^, worsened the R^2^, had variance inflation factors (VIF) greater than 2, or p>0.05. The final model was used to generate a raster of UFP concentration for the study domain, and it was evaluated using leave-one-area-out cross validation (LOAOCV): we split the study domain into four quadrants using the Watterson Expressway and Taylor Boulevard, and applied the model to each combination of three quadrants to evaluate model performance.

## Results and Discussion

### Ultrafine particle measurements

*Note: data in this section have not been deconvoluted*.

CP for the mobile MCPCs were 98% (average of all driving sessions), so data from one instrument was used for all further analyses. During all collocations, CP between the stationary and mobile MCPCs was <3% and Pearson’s ρ>0.99 (see Figure 9, Supplementary material), making scalar adjustments unnecessary.

The total length of drivable streets, excluding alleys but including streets by driving direction, in the study area was 340 km. Daytime, weekday UFP (Figure 2) ranged from 1,800-100,450 #/cm^3^. Levels in residential areas within 500 m of highways and major roads (arterials, primary collectors) ranged from 1,800-4,600 #/cm^3^, while levels proximate (<250 m) to or measured on these roads were in the uppermost quintile. The median UFP in the study domain was 6,850 #/cm^3^, which is slightly lower than levels reported from mobile measurements in the urban areas of Toronto, Canada (11,500 #/cm^3^), Norwich, UK (8,000 #/cm^3^), and Turin, Italy (14,000 #/cm^3^) but similar to urban background PNC in Pittsburg, PA (6,000-7,000 #/cm^3^) (6,11,13). The lower levels in this portion of Louisville could be attributable to the predominantly residential land use unlike the other investigations, which were city-wide and sampled from all land use types. It should also be noted that due to the absence of an established reference method for measuring PNC, studies use number concentrations for PM_0.1,_ PM_1_, or PM_2.5_ (as in this work) as proxies for UFP concentrations (6). Moreover, the particle detection efficiencies of CPCs at the lower cut points, which typically range from 1-10 nm, can greatly influence aggregated UFP levels because of the significant fraction of nucleation mode particles in vehicular exhaust (8).

**Figure 2.**
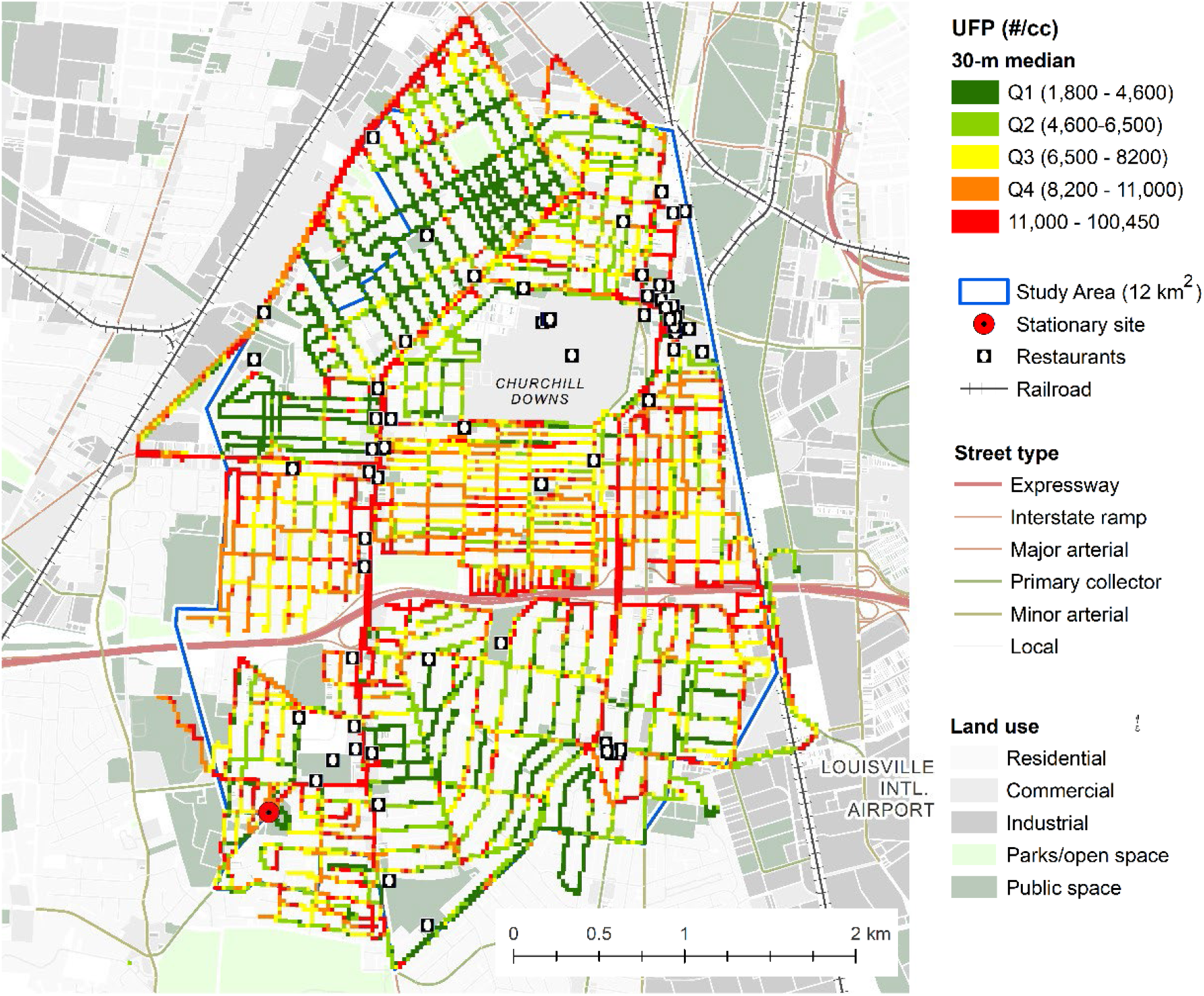
Daytime, weekday median UFP for 30-m road segments in the study domain; data collected in mobile campaigns between 2018-2020.

Expanding to nighttime and weekend data, we observed that median UFP was lowest during the daytime on weekdays (Figure 3). Mobile studies are typically not conducted at night, but assessments of stationary monitoring data corroborate the nighttime enhancement of UFP observed here, and this is consistent with diurnal patterns in the mixing layer depth (24). On the other hand, comparisons of PNC have found significantly higher levels during workdays (Mon-Fri) than non-workdays (Sat-Sun) (25,26). Morawska *et al*. measured lower median particle diameter on weekdays (44.2±0.3 nm) than on weekends (50.2±0.2 nm), thus concluding that the difference is dominantly driven by fresh motor vehicle emissions that contain particles of smaller diameters relative to those in the urban background (25). Consistent with lower vehicular emissions, variances were lower at both night and on weekends (on-road emissions cause instantaneous spikes in UFP; this was also observed by Giemsa *et al*. (27)), but I found the opposite pattern in median UFP, with weekend levels elevated compared to both daytime and nighttime levels on the weekdays. This is potentially due to the differing spatial scales in these assessments. While urban or regional scale UFP may decrease on weekends in response to lower traffic, changes in emissions due to local activity patterns may be enhanced at the neighborhood or residential scales (28).

**Figure 3.**
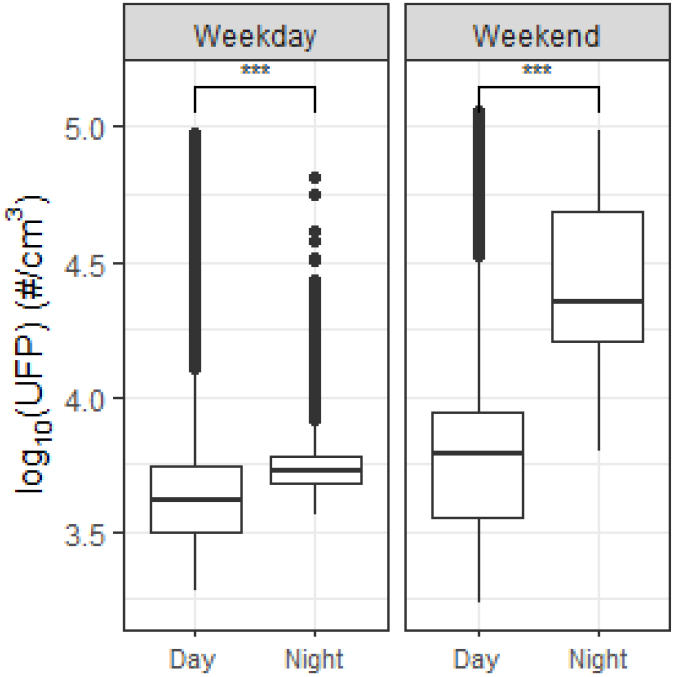
log_10_ median UFP collected on weekday (1:40 AM-3:20 AM, 2:45 PM – 5:30 PM, February 27, 2020) and weekend (2:00 – 5:45 AM, 3:20 PM – 7:30 PM, February 16, 2020) (all times in ET). Levels measured during day and nighttime were significantly different on both weekdays and weekends (Student’s t-test, ^***^ p<0.001).

Lastly, UFP enhancement near (<250 m) restaurants was higher than that from vehicular emissions near the interstate highway. This is illustrated in Figure 4: measurements from a weekday rush hour (3:50-6:00 PM ET, Friday; wind data in Figure 10, Supplementary material) segregated by distance from the interstate highway showed that relative to a residential (local) street 1 km upwind, median UFP was elevated by 25% on streets <250 m downwind of the highway; however, median UFP on streets 1 km and 1.5 km upwind of the highway but within 250 m of restaurants (9-10 establishments) was elevated by 165% and 200%, respectively. Numerous studies in the past decade have made similar observations of elevated particulate pollution in the vicinity of food service locations (18,21,29–33). Overall, this suggests that urban landscapes create persistent and hyperlocal hotspots, and that assessments of fine-scale environmental health effects may rely on short-term, mobile measurements to robustly characterize the resulting exposures.

**Figure 4.**
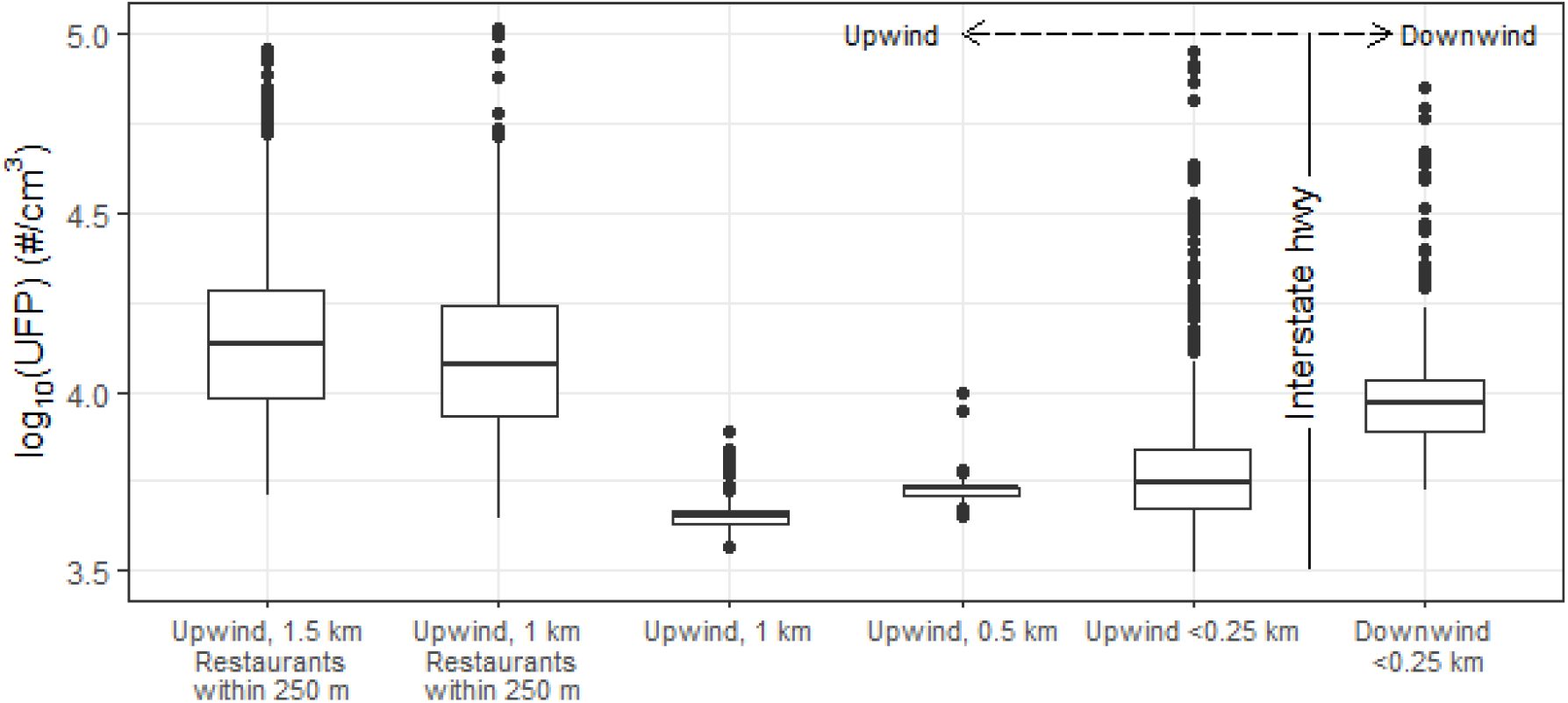
Distribution of median UFP per 30-m road segments relative to distance from an interstate highway, from daytime (3:50 – 6:00 PM ET), weekday mobile measurements (February 28, 2020).

### Deconvolution

The urban signal obtained from the splined minima of 3 min windows ranged from 3,860 #/cm^3^ to 11,750 #/cm^3^. These concentrations are comparable to the study domain-wide median (6,850 #/cm^3^) and are within the range of urban background levels (6,000 – 11,000 #/cm^3^) measured or estimated in other studies (6,11,13). A snippet of deconvoluted UFP from mobile monitoring, shown in Figure 5, demonstrates that this approach captured the diel pattern for these hours: weekday PNC, which reaches its daily maximum in mid-to late-morning, tapers off through the afternoon and early evening as the expanding mixing layer dilutes ground-level pollution (24,34,35). In this case, median UFP decreased from 10,400 #/cm^3^ to 6800 #/cm^3^ between 2:15 PM and 5:00 PM.

**Figure 5.**
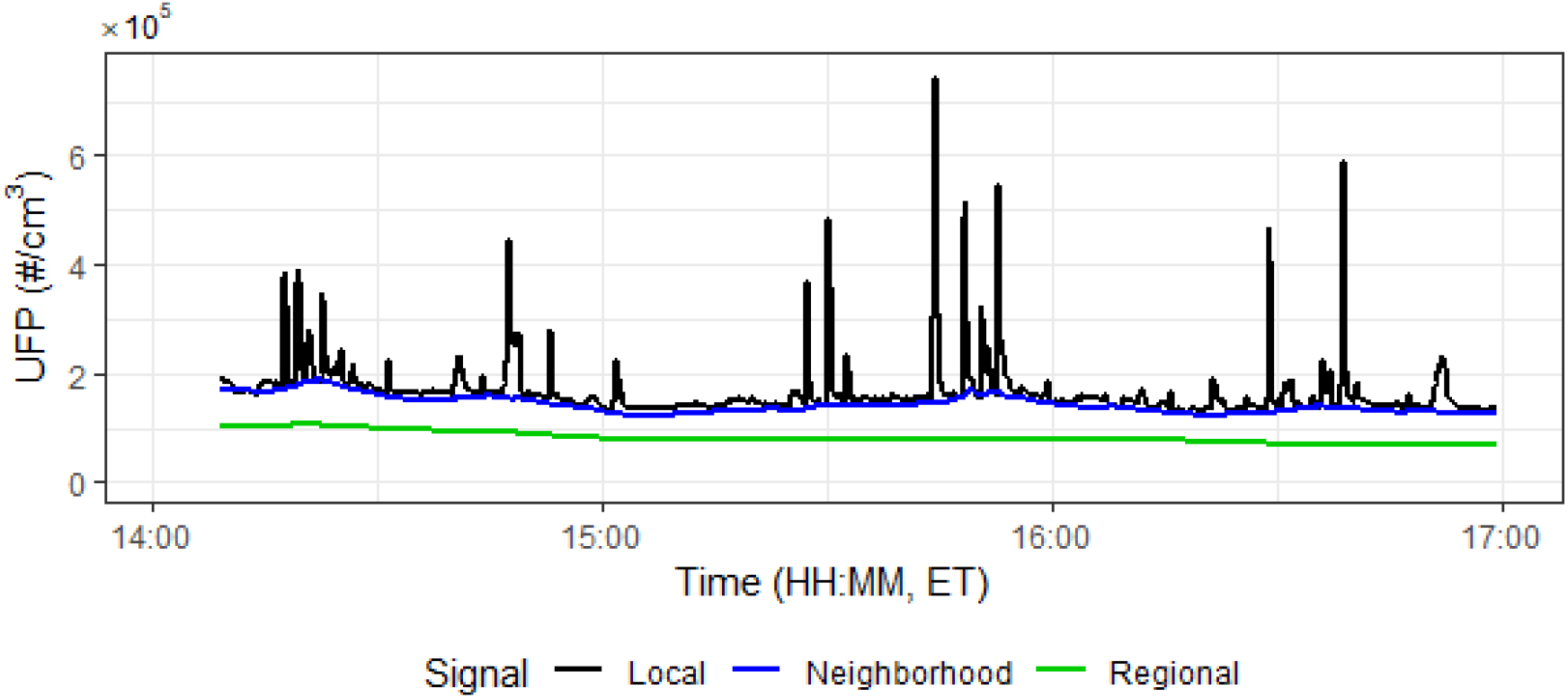
Local signal isolated from neighborhood (1-min minima) and regional (3-min minima) signals through median rolling deconvolution; data from mobile measurements on February 21, 2020.

Relative to the urban scale UFP, the contribution from the neighborhood scale ranged from 2,100-13,650 #/cm^3^. For the duration in Figure 5, the neighborhood-scale UFP was between 4,400 #/cm^3^ and 9,000 #/cm^3^. However, there was no discernible time-dependency in this signal. Rather, it likely reflected emissions from area-specific sources such as industrial activity and dispersed traffic. As shown in Figure 6, subtracting non-local signals revealed greater spatial heterogeneity in the measured 30-m median UFP (Fig 4.6 B) relative to no background standardization (Fig 6 A). Interestingly, the temporal scales suited to detect non-local signals in the Louisville study domain were much shorter (1 min and 3 min) than those identified in Toronto (8 minand 40 min) (11). While both studies relied on outlier-resistant median rolling methods, the broader scales for background interferences in Toronto are likely due to the larger spatial extent: compared to the 12 km^2^ region in the present work, the study in Toronto covered nearly 1,400 km^2^. In fact, removing 8 min and 40 min splined signals from measurements in Louisville drastically obscured small-area differences in median UFP (Fig 4.6 C). This suggests that background standardizations such as deconvolution, much like LUR modeling, must be conducted empirically, but are highly effective in evincing hyperlocal heterogeneity in UFP.

**Figure 6.**
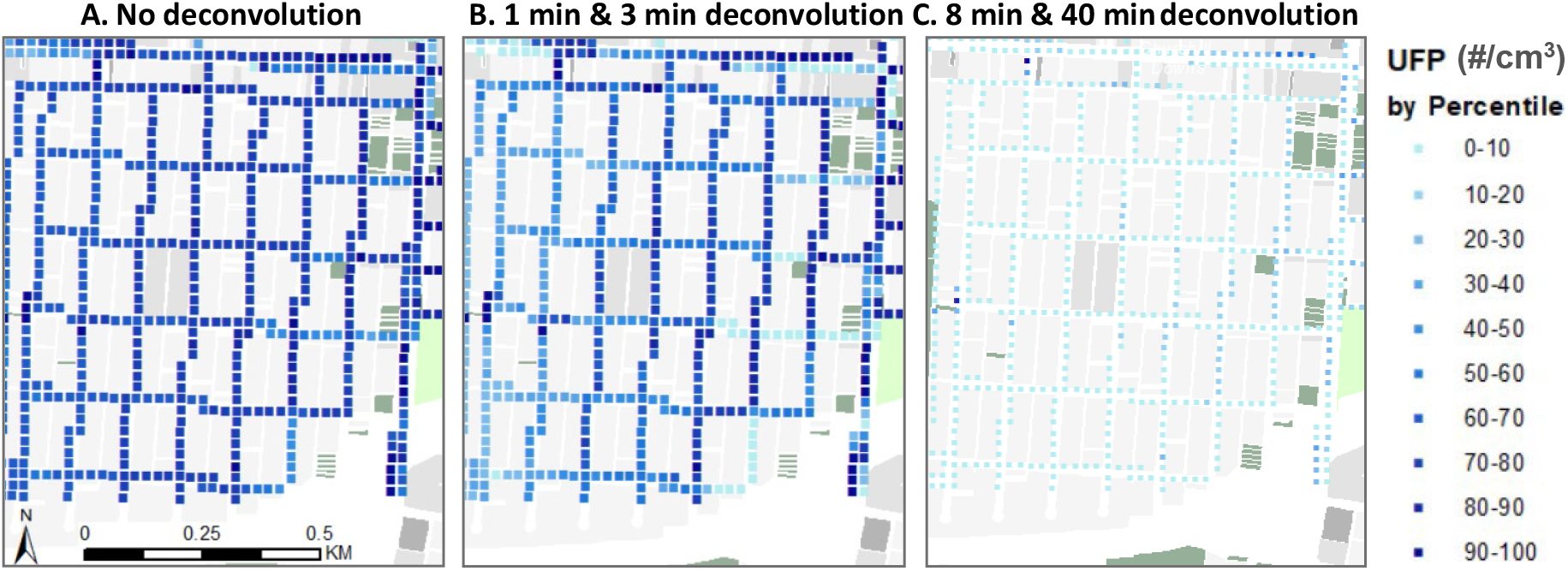
Spatial heterogeneity in 30-m median UFP with no deconvolution (A), 1 min and 3 min rolling minima subtracted (determined in this study) (B), and 8 min and 40 min rolling minima subtracted (from Shairsingh *et al*. (11)) (C).

### Modeling results

The final regression model explained 61% of the variance in deconvoluted UFP. Variables quantifying proximity to roadways and restaurants were the most correlated with UFP and thus, considered first. Then, parameters denoting cumulative occurrence of greenness (NDVI) and industrial land use were added, but interestingly, most area metrics did not improve the adjusted R^2^. The final model (Table 1) consisted of 13 predictors, all with VIF < 2; Figure 7 shows the resulting surface of UFP concentration.

**Table 1.** Summary of multivariable regression model for the deconvoluted, 30-m log(UFP) with standardized coefficients

| Type | Predictor | Coefficient | Standard error | t | Prob>t | VIF |
| --- | --- | --- | --- | --- | --- | --- |
| **ln(UFP) (adj. R<sup>2</sup> = 0.61, p<0.001)** |  |  |  |  |  |  |
|  | Constant | 7.88x10<sup>-3</sup> | 7.57x10<sup>-3</sup> | 1.03 | 0.302 |  |
| Total length | Highway (<100 m) | 2.84x10<sup>-1</sup> | 7.00x10<sup>-3</sup> | 4.06 | 0.000 | 1.051 |
|  | Primary collector (<250 m) | 3.96x10<sup>-2</sup> | 6.96x10<sup>-3</sup> | 5.69 | 0.000 | 1.032 |
|  | Major arterial (<250 m) | 3.09x10<sup>-2</sup> | 7.02x10<sup>-3</sup> | 4.40 | 0.000 | 1.053 |
| Area | NDVI (500 m) | -4.20x10<sup>-3</sup> | 7.09x10<sup>-4</sup> | -5.78 | 0.000 | 1.131 |
|  | Leaf biomass (<200 m) | -1.01x10<sup>-3</sup> | 9.38 x10<sup>-4</sup> | -6.32 | 0.000 | 1.125 |
|  | Leaf biomass (<300 m) | -1.98x10<sup>-3</sup> | 9.94 x10<sup>-4</sup> | -6.86 | 0.000 | 1.169 |
| Count | Traffic signals (<150 m) | 2.24x10<sup>-2</sup> | 6.96x10<sup>-3</sup> | 3.53 | 0.000 | 1.027 |
|  | Restaurants (<350 m) | 1.41x10<sup>-2</sup> | 5.31x10<sup>-3</sup> | 2.69 | 0.007 | 1.013 |
| Proximity | Commercial | 2.45x10<sup>-2</sup> | 6.98x10<sup>-3</sup> | 3.49 | 0.000 | 1.043 |
|  | Residential | -1.24x10<sup>-2</sup> | 6.98x10<sup>-3</sup> | -1.99 | 0.047 | 1.034 |
|  | Industrial | 1.76x10<sup>-2</sup> | 6.94x10<sup>-3</sup> | 2.54 | 0.010 | 1.022 |
|  | Restaurants | 1.28x10<sup>-2</sup> | 5.29x10<sup>-3</sup> | 2.45 | 0.015 | 1.120 |

**Figure 7.**
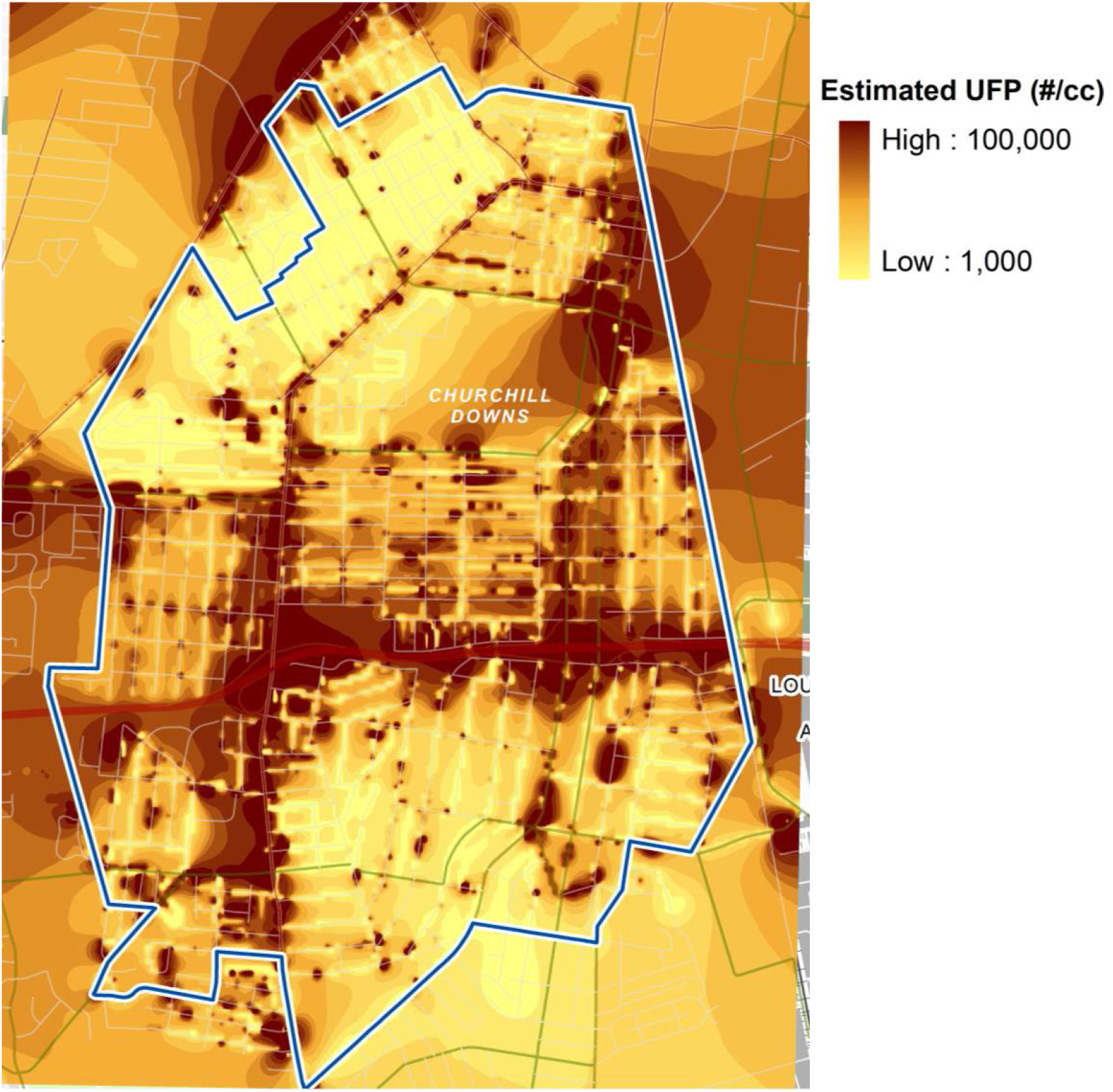
Modeled 30-m UFP concentration for the study domain

Four explanatory variables quantified cumulative occurrence or proximity to previously known sources of elevated UFP—vehicular traffic and restaurant cooking. In the case of restaurants, the distance to the nearest location and the count within a 350 m buffer were significant predictors. This buffer distance is consistent with the zone of elevated UFP noted earlier (Figure 4) and with the exponential decay distance in organic aerosol—albeit by mass concentration—reported in a previous study, where the steepest decline was observed within 300 m downwind of restaurants (18).

The only predictors negatively associated with UFP were proximity to residential land use and greenness by cumulative occurrence (NDVI within 500 m, leaf biomass within 200 m and 300 m), a unit increase in which corresponded to modest UFP decreases between 0.1-1.2%. It is unclear whether this decrease is due to mitigation by vegetation or due to the absence of emission sources: though the final predictors exhibited low collinearity (VIF<2) overall, it is possible that the presence of open spaces may be inversely correlated with the presence of roadways in small portions of the study domain. Limited number of LUR models have fitted UFP from mobile measurements with NDVI metrics; rather, most studies include parameters of open space or parks. Such variables were not retained in this study, but Hankey *et al*. reported negative associations between particle number concentrations and open spaces within 25 m and 1,500 m, and Xing *et al*. showed that urban vegetation removed at most 1% of pollutants through deposition (10,36). The associations between UFP and greenness parameters detected in this study support Xing *et al*.’s assertion that removal through deposition, which varies with leaf area, has limited impact on variability in pollutant concentrations; rather, leaf biomass—a proxy for foliage—alters pollutant concentrations in microenvironments through mechanical interference.

LOAOCV revealed that the model overpredicted UFP in the upper left and lower right quadrants of the study domain, which extend farther from the interstate highway and have food service locations mostly along the perimeter, by 9-13%. In contrast, the model underpredicted UFP in the upper right quadrant, which is transected by several arterial roads and embedded with numerous restaurants. This suggests that the model can be refined if fitted to measured data with more repetitions in each section. Moreover, since deconvolution can be applied directly to mobile measurements (i.e. determining spline windows using mobile-platform data rather than the stationary-site data), all three instruments could be simultaneously deployed for mobile monitoring, thus expanding the spatial coverage of each session.

The predictive capability of this model is consistent with previous assessments using 20 or fewer repeated measurements along the same route (10,37). Studies have found that LUR models require longer-term, seasonally robust UFP estimates from mobile measurements to obtain performance better than 0.5< adjusted R^2^ <0.6. This is certainly recommended for future analyses in the Louisville domain. Though the deconvolution method used here does not necessitate season- and year-specific monitoring to identify the true background, monitoring that exceeds 10-20 drives per season will better characterize long term patterns in local UFP.

On a final note, another approach to improve the variance explained by the model is to include additional predictors. While land use, land cover, and traffic-related parameters were considered extensively in this work, variables for local meteorology were not included. Numerous studies have reported significant associations between deconvoluted UFP and meteorological variables (wind speed, wind direction (in 30-45° sectors), relative humidity) in LUR models (11,12,38,39). Wind velocity, in particular, deserves further discussion as two common strategies to mitigate pollution exposure—sound walls and green interventions—interfere with wind patterns at spatial scales in which the present work and previous investigations have shown persistent UFP patterns (19,21). Unlike in these two assessments of coastal cities, however, wind direction in Louisville is heterogeneous over the temporal extent of the mobile measurements (see Figure 11, Supplementary material).

The strong downwind signals from highways observed in this work and extensively explored in others compel the consideration of time-matched wind direction as a predictor. Indeed, if the driving speed of the mobile platform—typically 8-30 kph—is used to calculate bin widths for rolling minima splines to effectively detect background signals, it is reasonable to speculate that wind speed—which ranges from 6-30 kph for light to moderate breezes—can also be a determinant of hyperlocal UFP (40). In fact, wind speeds have been used to determine temporal scales to deconvolute PNC (41). It is important to note, however, that wind speeds in urban microenvironments exhibit micrometeorological behavior; features of the built and natural environment, including sound walls and tree canopies, may stagnate wind flow or generate turbulence (42). In other words, hyperlocal wind velocity may be just as dependent on land use parameters as pollutant levels. LUR models that found statistically significant associations between wind speed or direction with UFP have been developed for coastal or lakeside cities such as Toronto, Vancouver, and Boston (11,12,43,44) or in assessments based on fixed-site monitoring (40). In land areas adjacent to major water bodies, sea or lake breezes maintain persistent wind patterns that may be retained hyperlocally; this is likely not the case in inland cities such as Louisville (45).

## Conclusion

This study mapped and modeled 30-m UFP in a section of Louisville, KY, demonstrating that mobile monitoring strategies developed by assessments of larger-scale urban air quality characterizations can evince hyperlocal pollution patterns at even finer spatiotemporal scales. The model was fitted to temporally deconvoluted UFP such that regional and neighborhood scale baselines were removed to better capture spatial heterogeneity associated with local land use parameters. An implication of this work is that, given the dependence of UFP on characteristics of the built and natural environment, rational implementation of mitigation strategies may alleviate hyperlocal UFP exposures.

## Data Availability

All data produced in the present study are available upon reasonable request to the authors

## Supplementary material

**Figure 8.**
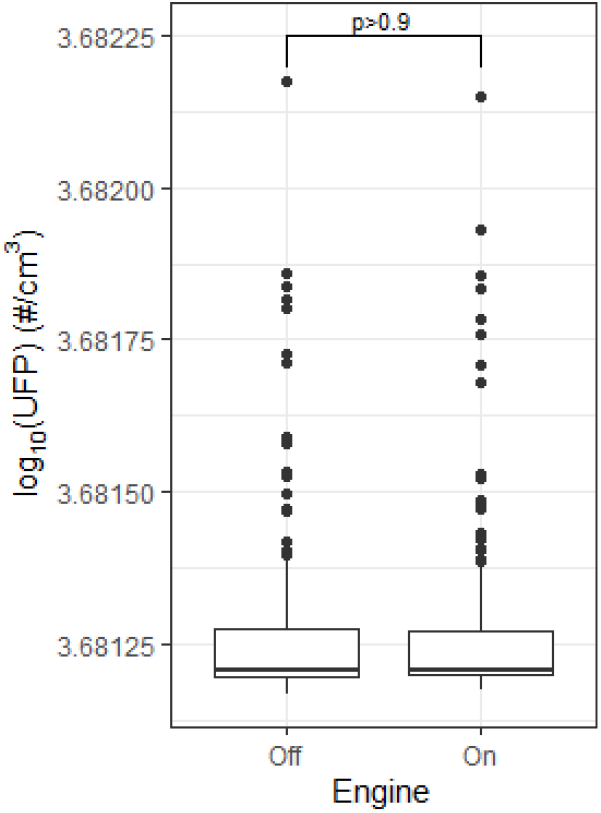
1-second log_10_(UFP) measured on the gasoline-powered mobile platform with the engine off and on (20 min each).

**Figure 9.**
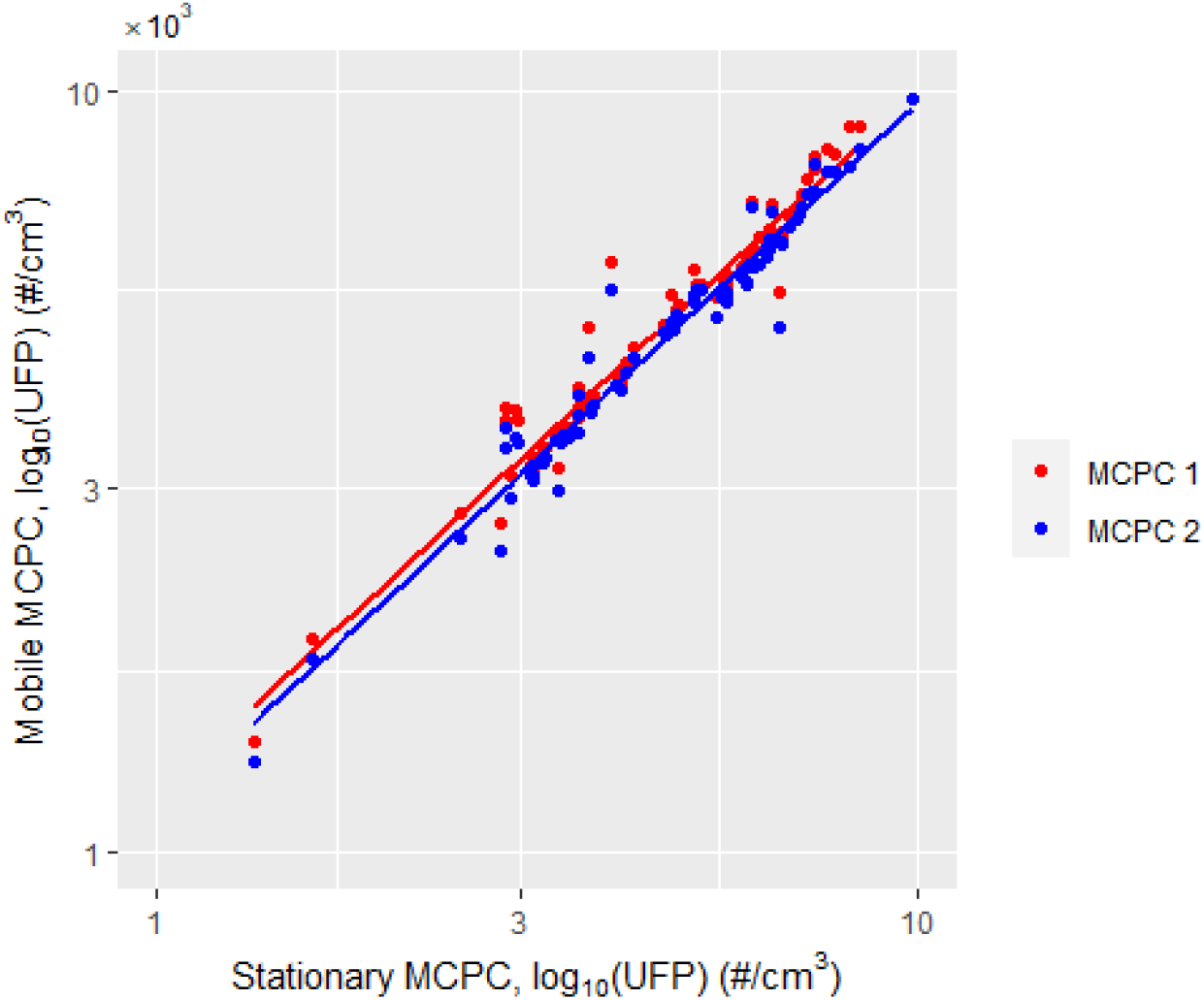
Collocated (mobile platform ∼10 m from stationary site) 1-second UFP measured by the stationary MCPC and the mobile MCPCs (MCPC 1 and MCPC 2); Pearson’s correlation coefficients were ρ_MCPC 1_ = 0.9978 (95% CI 0.9965, 0.9973) and ρ_MCPC 2_ = 0.9968 (95% CI 0.9963, 0.9972).

**Figure 10.**
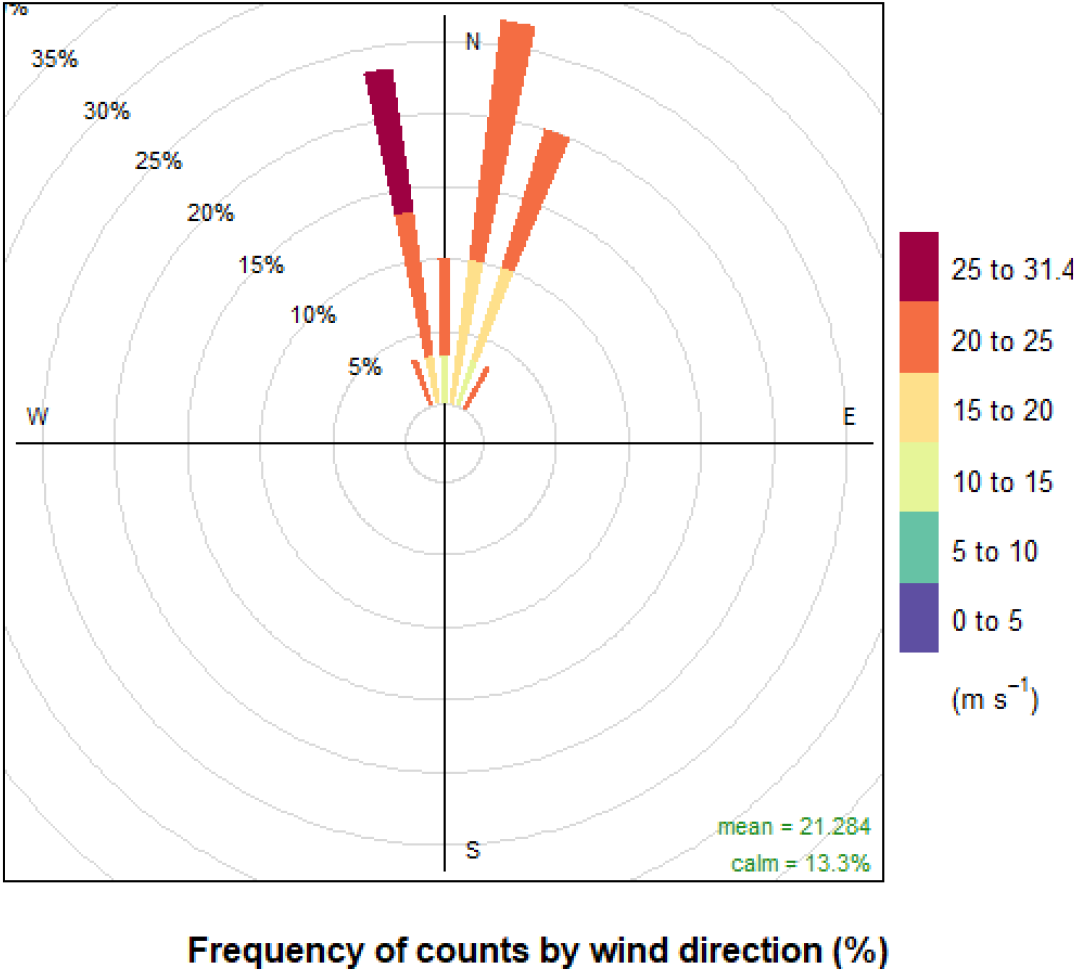
5-min wind for the measurement duration over which data for Figure 4.4 was collected (3:50 – 6:00 PM ET, February 28, 2020); data obtained from Kentucky ASOS for Louisville International Airport (https://mesonet.agron.iastate.edu/request/download.phtml)

**Figure 11.**
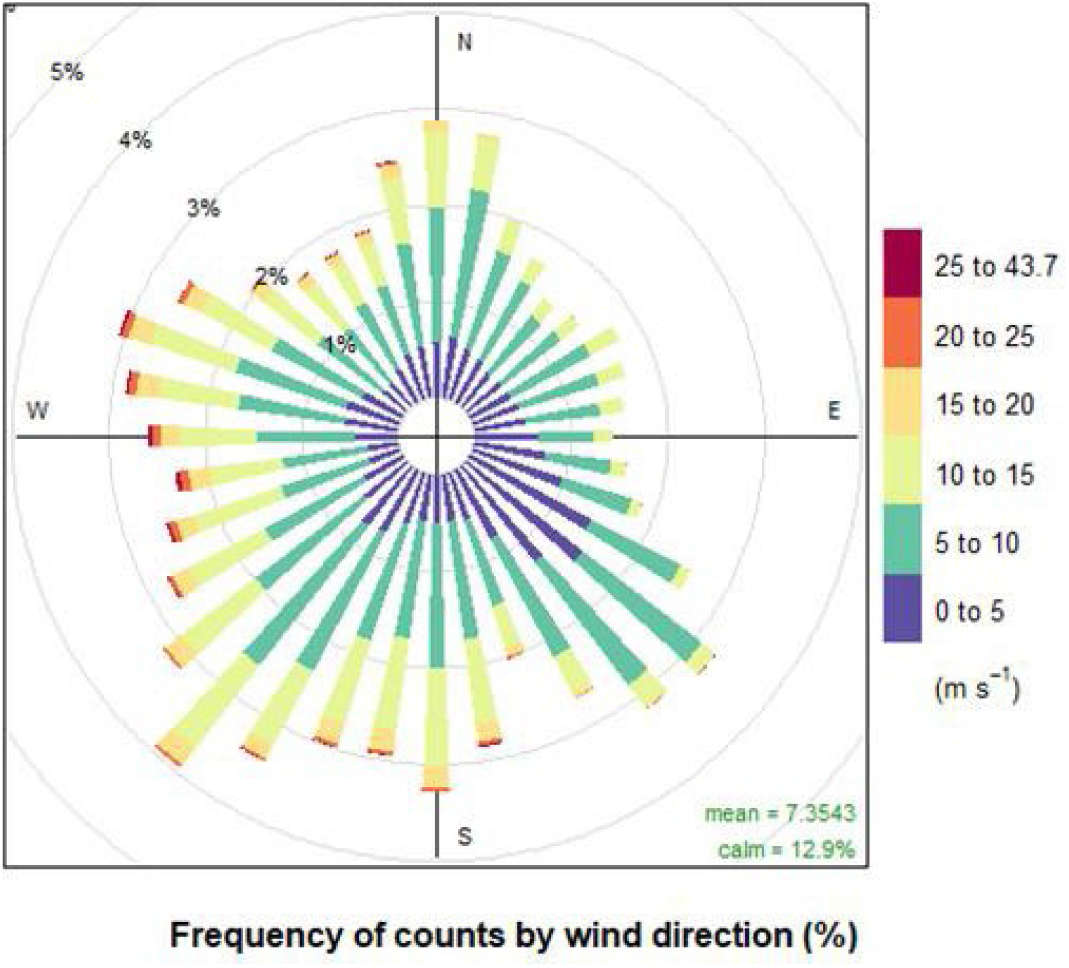
5-min wind for 2019-20 as measured at Louisville International airport; data obtained from Kentucky ASOS (https://mesonet.agron.iastate.edu/request/download.phtml)

